# Modelling COVID-19 mutant dynamics: understanding the interplay between viral evolution and disease transmission dynamics

**DOI:** 10.1101/2024.06.04.24308411

**Authors:** Fernando Saldaña, Nico Stollenwerk, Maíra Aguiar

**Affiliations:** Basque Center for Applied Mathematics (BCAM), Bilbao, Spain; Ikerbasque, Basque Foundation for Science, Bilbao, Spain

## Abstract

Understanding virus mutations is critical for shaping public health interventions. These mutations lead to complex multi-strain dynamics often underrepresented in models. Aiming to understand the factors influencing variants’ fitness and evolution, we explore several scenarios of virus spreading to gain qualitative insight into the factors dictating which variants ultimately predominate at the population level. To this end, we propose a two-strain stochastic model that accounts for asymptomatic transmission, mutations, and the possibility of disease import. We find that variants with milder symptoms are likely to spread faster than those with severe symptoms. This is because severe variants can prompt affected individuals to seek medical help earlier, potentially leading to quicker identification and isolation of cases. However, milder or asymptomatic cases may spread more widely, making it harder to control the spread. Therefore, increased transmissibility of milder variants can still result in higher hospitalizations and fatalities due to widespread infection. The proposed model highlights the interplay between viral evolution and transmission dynamics. Offering a nuanced view of factors influencing variant spread, the model provides a foundation for further investigation into mitigating strategies and public health interventions.

## 1 Introduction

The success of SARS-CoV-2 as a global pandemic can be largely attributed to its significant capacity for genetic mutation, enabling rapid adaptation to various conditions, host populations, and modes of transmission [1]. This mutation process leads to the emergence of new variants that may exhibit changes in key characteristics like transmissibility, virulence, or resistance to treatments and immune responses. Currently, global genomic sequencing and surveillance efforts have identified numerous variants of SARS-CoV-2, with only a small fraction categorized as Variants of Concern (VOCs) due to their potential public health implications [2]. Notably, variants such as Delta and Omicron have attracted attention for their increased transmissibility and ability to evade immunity. The heightened transmissibility of variants accelerated the spread of the virus, overwhelming healthcare infrastructure. Furthermore, the VOCs’ capacity to evade acquired immunity from prior infection or vaccination raised concerns about the effectiveness of existing control measures. The rise of VOCs has presented substantial obstacles to public health managing virus transmission and reducing the burden on healthcare systems [3, 4].

Numerous modelling studies have been conducted to explore SARS-CoV-2 dynamics and guide public health strategies during the pandemic [5, 6]. However, most modelling efforts have focused on singular variants and only a relatively low number of studies has investigated multi-strain SARS-CoV-2 dynamics [2, 7, 8, 9, 10]. To effectively address SARS-CoV-2 variants, an integrated strategy is needed, including enhanced surveillance, robust vaccination efforts, and targeted public health interventions. Furthermore, to design successful interventions, it is essential to understand the drivers of viral fitness and evolution. These factors include the speed of viral replication in host cells and its capacity for efficient transmission between individuals [3].

For SARS-CoV-2 and other viruses with relatively short infectious periods, transmissibility is argued to be the primary driver of evolution, especially in immunologically naive hosts [4]. One of the key quantities used to quantify the transmissibility of a virus is its basic reproduction number *R*_0_ [11, 6, 12]. A virus can maximize its *R*_0_ by increasing its intrinsic transmissibility, such as enhancing viral shedding, survival outside the host, and the ability to establish infection in new hosts. Viruses that can maintain infectiousness over longer periods may have a better chance of persisting in a population and establishing ongoing transmission chains. Therefore, in addition to increasing intrinsic transmissibility, a virus can increase its *R*_0_ by extending its duration of infectiousness [4]. In this study, we develop a parsimonious two-strain stochastic *SHAR* model accounting for multiple factors, including asymptomatic transmission, mutations, spillover events, and the potential importation of diseases from external sources into the population of interest. The inclusion of asymptomatic transmission acknowledges the role of individuals who may unknowingly transmit the virus, contributing to its spread within the population. Additionally, incorporating mutations and/or consideration of spillover events are essential for understanding the risks of the emergence of novel variants and their potential to cause outbreaks [13, 14, 15]. Finally, by accounting for the possibility of disease importation from outside the population of interest, our model allows us to evaluate how infectious diseases can be introduced into a population through travel and to quantify their impact on transmission patterns and outbreak dynamics. Instead of providing quantitative predictions of disease spread, here we explore various possibilities to gain qualitative insights into the factors influencing which variants prevail at the population level.

## 2 Methods and results

The modelling framework used in this study is a two-strain stochastic epidemic model that builds upon the classical *SIR* (susceptible-infectious-recovered) model. Three significant extensions are taken into account. First, the infectious class *I* is divided into severe infections prone to hospitalization (*H*) and individuals with mild or asymptomatic infection (*A*). Second, the model incorporates the strain structure of pathogens, considering a two-strain framework that accounts for both a wild-type strain of the virus and a mutant strain resulting from genetic mutations. Third, an import factor *ρ* is considered to mimic the possibility of susceptible individuals acquiring the infection via an undetected infection chain that started outside the population of interest. The parameter *ρ* can also account for zoonotic spillover events in which animal reservoirs transmit the infection to a human [16]. To properly explain the features of the framework, we first describe the mean-field version of the *SHAR* (susceptible-hospitalized-asymptomatic-recovered) model that serves as a basis for the general two-strain stochastic model. Then we formulate the two-strain deterministic *SHAR* model followed by its stochastic counterpart.

### 2.1 The deterministic SHAR model with import

Besides the transmission rate *β*, the recovery rate *γ* and the waning immunity rate *α* presented in the usual *SIR* model, the *SHAR* framework incorporates two additional parameters: the severity ratio *η* represents the fraction of infected individuals who develop severe symptoms requiring hospitalization, and a transmission enhancement factor *ϕ* which differentiates the infectivity of individuals in the asymptomatic (*A*) class from the baseline infectivity *β* of the hospitalized (*H*) class.

Finally, incorporating the influence of import *ρ* on disease transmission within the population, which reflects the potential introduction of infections from external sources, the *SHAR* model is formulated as follows:

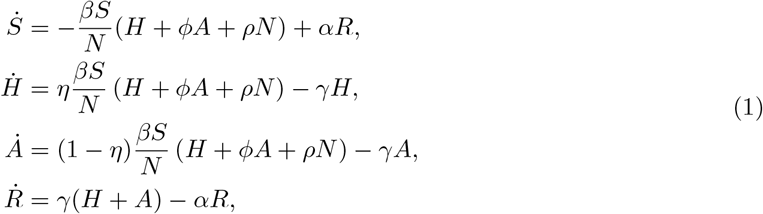

where the total population size *N* = *S* + *H* + *A* + *R* is a positive constant. Observe that with a positive import *ρ* > 0, the model (1) does not admit a disease-free equilibrium. Still, for low values of *ρ* (around the order of 10^−5^) numerical simulations show a positive equilibrium very close to zero in the subcritical regime (see Figures 2 and 3). Setting the right-hand side of model (1) equal to zero, a direct computation shows that system (1) has a unique endemic equilibrium given by

**Figure 1.**
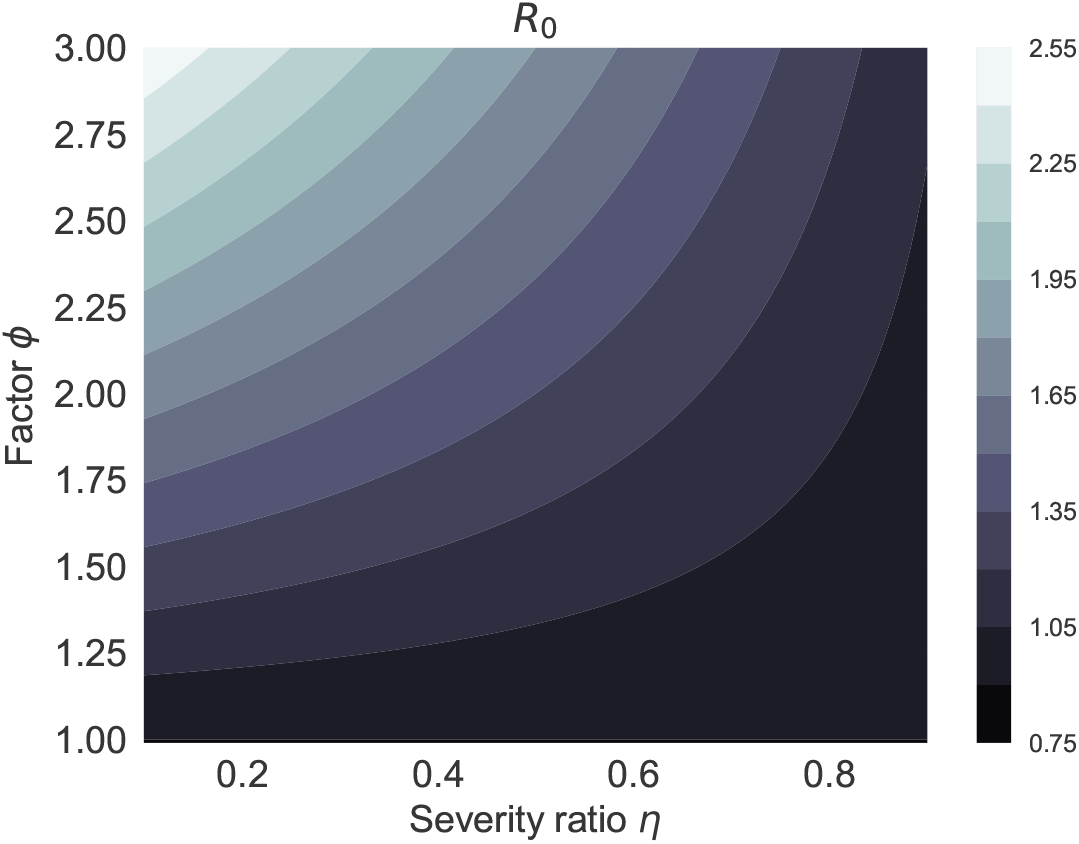
Contour plot for the basic reproduction number *R*_0_ (3) as a function of the hospitalized fraction *η* and the factor *ϕ*. Other parameters are fixed as *γ* = 1*/*7, *β* = 0.9*γ*.

**Figure 2.**
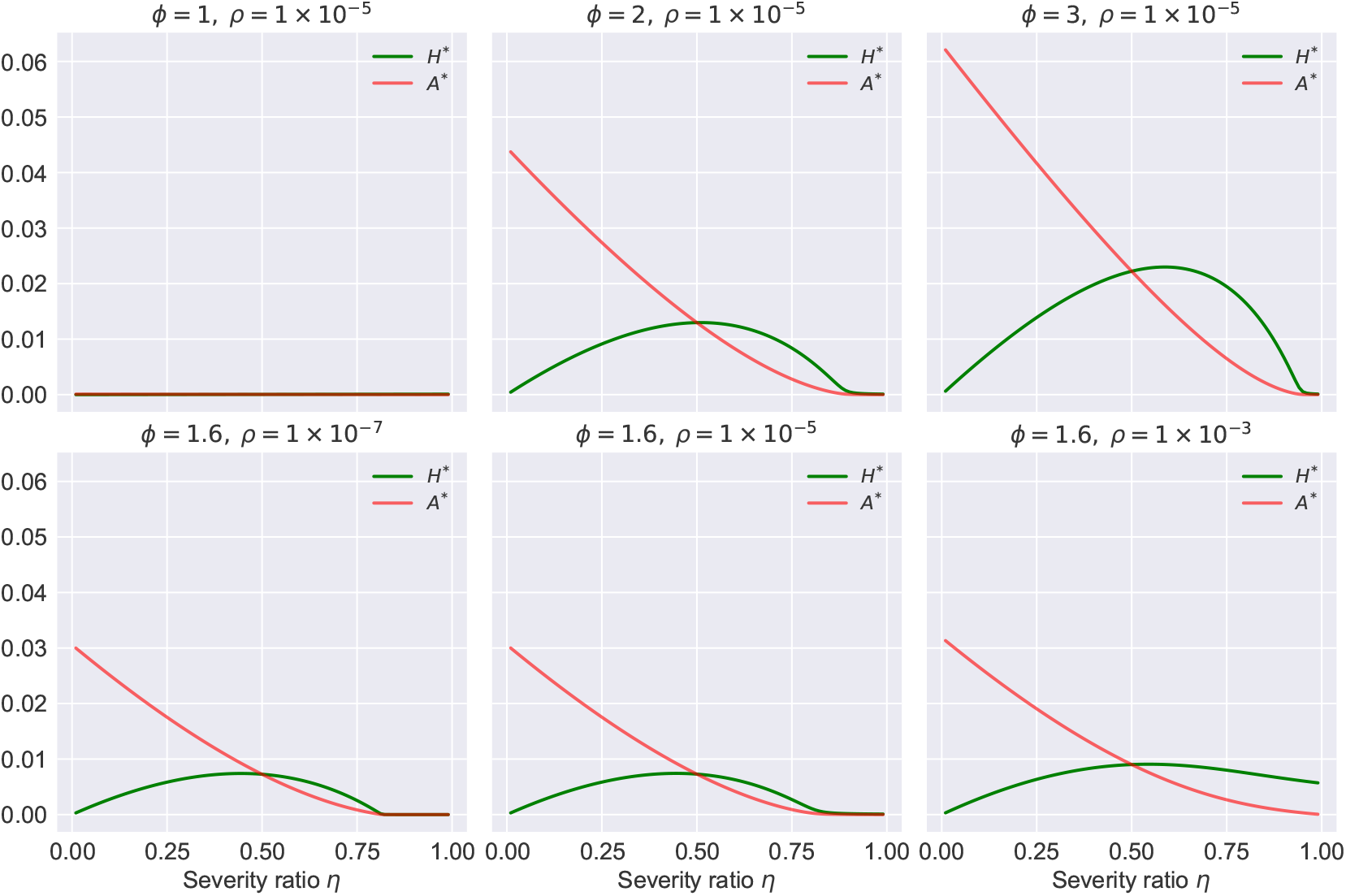
Endemic equilibria *H*^∗^ and *A*^∗^ of model (1) as a function of *η* for different values of *ϕ* and the import *ρ*. Other parameters are fixed as *γ* = 1*/*20, *β* = 0.9*γ, α* = 1*/*180, *N* = 1.

**Figure 3.**
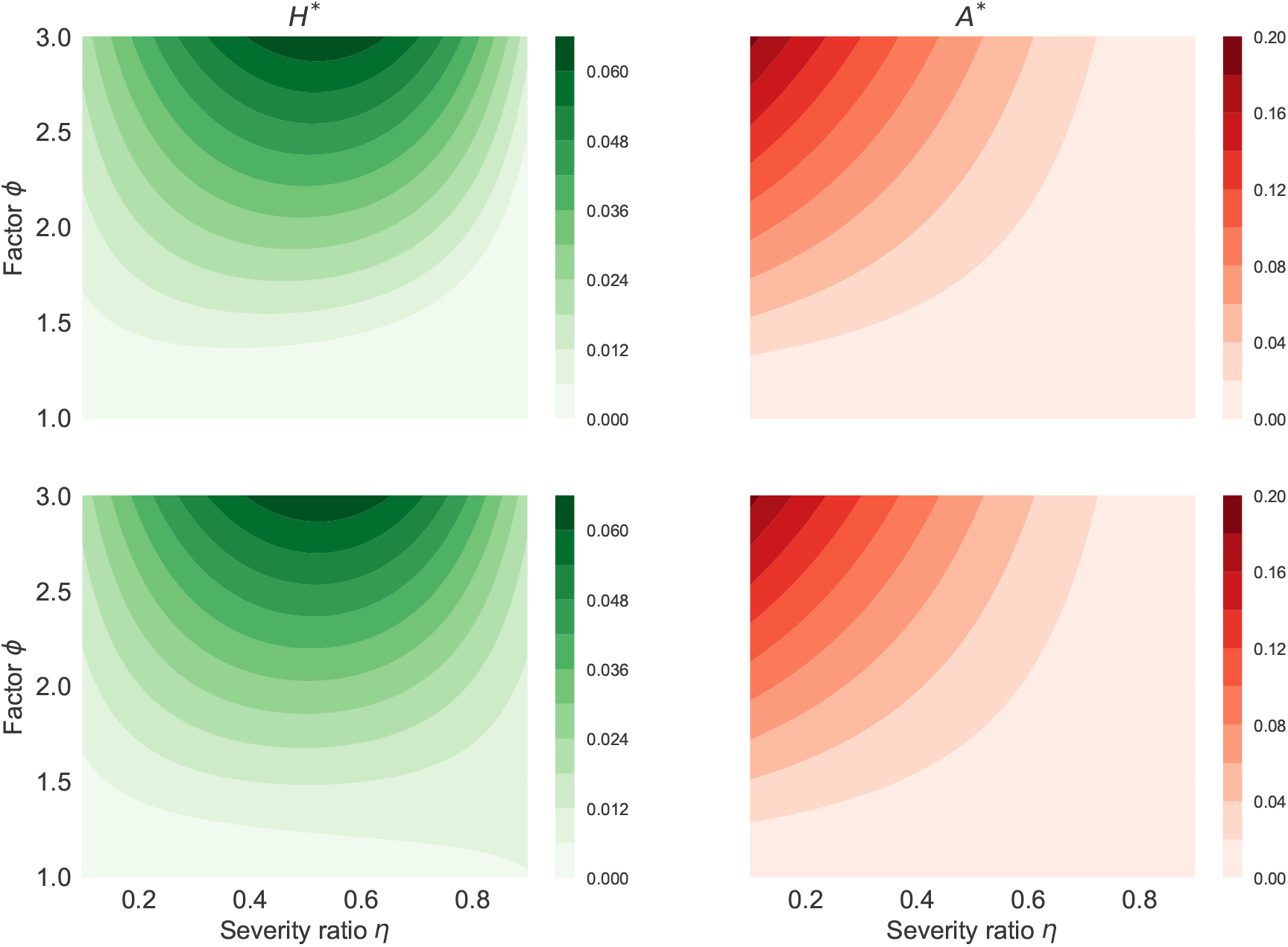
Contour plot for the endemic equilibria *H*^∗^ and *A*^∗^ of model (1) as a function of *η* and *ϕ*. The value of the import *ρ* is 1×10^−6^, and 1×10^−3^, for the first and second row, respectively. The rest of parameters in the one-strain SHAR model (1) are fixed as *γ* = 1*/*20, *β* = 0.9*γ, α* = 1*/*180, *N* = 1.

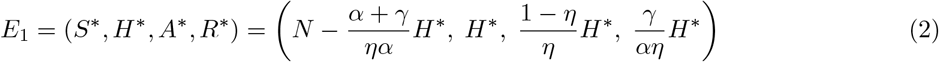

The hospitalized class at equilibrium *H*^∗^ is given by the solution of the following second-order polynomial

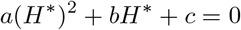

Where

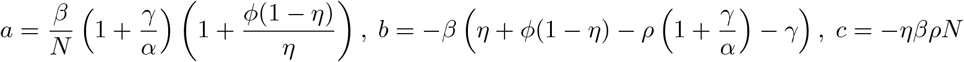

Observe that *a* > 0 and *c* < 0, hence 4*ac* > 0 and 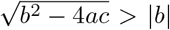 , therefore the only biologically feasible equilibrium is

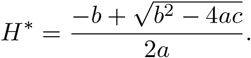

The basic reproduction number for the model (1) can be straightforwardly obtained using the next generation method [11] as

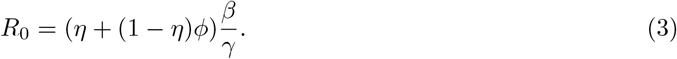

Observe that the basic reproduction number *R*_0_ is the weighted average of the secondary infections caused by the *H* class *R*_0,*H*_ = *β/γ* and the number of secondary infections caused by the *A* class *R*_0,*A*_ = *ϕβ/γ*, so *R*_0_ = *ηR*_0,*H*_ + (1 − *η*)*R*_0,*A*_.

If *ρ* = 0, that is, in the absence of import, the constant *c* is zero and the equilibria of the *SHAR* model are determined by the solutions of *a*(*H*^∗^)^2^ + *bH*^∗^ = 0. Clearly, one solution is *H*^∗^ = 0 so the disease-free equilibrium is *E*_0_ = (*N*, 0, 0, 0). The second solution i.e the disease endemic equilibrium becomes

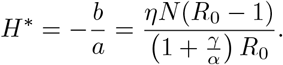

Hence, for *ρ* = 0, a positive endemic equilibrium only exists if *R*_0_ > 1. The stability of the disease-free equilibrium *E*_0_ can be obtained using the Jacobian matrix *J* of the *SHAR* model. Considering that the population size is constant, we can exclude the equation for *S* and obtain the following expression for the Jacobian evaluated at *E*_0_:

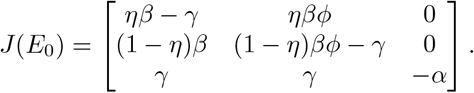

The Jacobian matrix has an eigenvalue *λ*_1_ = −*α*, and the other two eigenvalues are determined by eigenvalues of the 2 × 2 sub-matrix

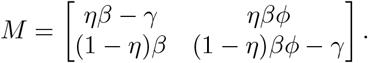

Observe that if *R*_0_ < 1 then (*η* + (1 − *η*)*ϕ*)*β* < *γ*. Hence, the determinant of *M* given by *det*(*M* ) = *γ*(*γ* − (*η* + (1 − *η*)*ϕ*)*β*) satisfies *det*(*M* ) > 0 if and only if *R*_0_ < 1. Likewise, the trace of *M* given by *tr*(*M* ) = (*η* + (1 − *η*)*ϕ*)*β* − 2*γ* satisfies *tr*(*M* ) < 0 if and only if *R*_0_ < 1. This implies that the eigenvalues of *M* are negative if and only if *R*_0_ < 1 and therefore *E*_0_ is locally asymptotically stable if *R*_0_ < 1 and unstable if *R*_0_ > 1.

It is important to remark that the transmission rate *β* can be decomposed as a product *β* = *kq*, where *k* is the average number of contacts per person per unit of time and *q* is the probability of transmission given a contact between a susceptible and an infectious individual [17]. In the context of COVID-19, there has been some discussion about the transmission potential of asymptomatic carriers. Some studies suggest that asymptomatic people have usually a lower viral load when compared with symptomatic people and hence are less likely to transmit the virus [18, 19]. However, this only implies that the transmission probability *q* is lower for asymptomatic individuals than for symptomaticindividuals. Several other studies have argued that asymptomatic carriers are more prone to have social interactions and therefore a higher contact rate *k* than people with severe symptoms [20, 21, 22, 23, 24, 25, 26, 10, 27]. Consistent with these studies, here we assume that the factor *ϕ* in the model (1) is equal to or greater than one. The fraction 0 < *η* < 1 of infected individuals that develop severe disease is also a critical parameter in the basic reproduction number (3). Figure 1 shows the basic reproduction number (3) as a function of *η* and *ϕ*. Observe that although it is assumed that *β* = 0.9*γ* so the *SIR* sub-system is below criticality, considering *ϕ* > 1 allows the *SHAR* model to reach a supercritical regime with *R*_0_ > 1. Figure 2 depicts the endemic equilibria *H*^∗^ and *A*^∗^ of model (1) as a function of *η* for different values of *ϕ* and the import factor *ρ*. For the first row in Figure 2, *ϕ* takes the values 1, 2, and 3 for the first, second, and third columns, respectively. For the second row in Figure 2, the value of the import factor *ρ* is 1×10^−7^, 1×10^−5^, and 1×10^−3^, for the first, second and third column, respectively. Observe that the endemic equilibrium increases significantly as a function of *ϕ*. On the other hand, the impact of *ρ* on the endemic equilibrium is mainly visible when the system is below or close to criticality (*R*_0_*≤*1). These results can be observed more clearly in Figure 3 which shows a contour plot for the hospitalized and asymptomatic infectious classes at the endemic equilibrium as a function of *η* and *ϕ* for different values of the import factor *ρ*. Again, considering *ϕ* > 1, the *SHAR* model can have a positive endemic equilibrium. Observe (see Figure 3) that for *ϕ* close to 1 and low import factor the endemic equilibrium is very close to zero but considering *ϕ* > 1 or a higher value for *ρ* the area of such region decreases significantly.

### 2.2 The two-strain deterministic extension of the SHAR model with import

We now extend the *SHAR* model described in section 2.1 by incorporating strain structure into the system. Assuming that the wild-type strain of the virus (denoted by subscript *w*) undergoes mutations at a rate *ϵ*, leading to the appearance of a mutant strain (denoted by subscript *m*). The transmission rate, the severity ratio, and the recovery rate are strain-dependent. For simplicity, it is assumed that individuals in the recovered class *R* are immune to both strains. However, this immunity is not long-lasting, and recovered individuals lose their immunity at a rate *α*. The total population (*N* ) is the sum of the six mutually exclusive epidemiological classes of the model, i.e., *N* = *S* +*H*_*w*_ +*A*_*w*_ +*H*_*m*_+*A*_*m*_+*R*. The model dynamics for the two-strain deterministic *SHAR* model are described by the following system of differential equations:

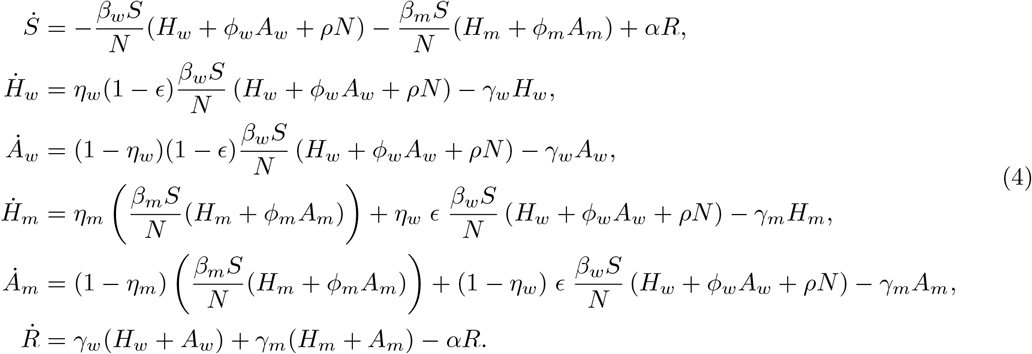

Observe that, as before, if the import is *ρ* = 0, then at the disease-free equilibrium *S* = *N* and the other variables in system (4) are equal to zero. If *ρ* > 0 there is no disease-free equilibrium. Due to the high nonlinearity of the two-strain SHAR model (4) it is no longer possible to obtain an analytical expression for the endemic equilibria as we did for the one-strain SHAR model. Hence, to understand the long-term dynamics of the two-strain model (4) we perform a global sensitivity analysis (GSA). Since the values for several important parameters in the model (4) such as the disease-severity ratios (*η*_*w*_, *η*_*m*_), the change of baseline infectivities (*ϕ*_*w*_, *ϕ*_*m*_), the mutation rate (*ϵ*), the import factor (*ρ*), and so on, present considerable uncertainty or at least wide ranges, the GSA becomes a useful tool to measure the individual importance of each parameter as well as their joint effect on model outcomes. To perform the GSA, we use Sobol’s method which is based on variance decomposition techniques and provides a quantitative measure of the contributions of the input parameters to the output variance [28]. Although there are several types of GSA (e.g. weighted average of local sensitivity analysis, partial rank correlation coefficient, multiparametric sensitivity analysis), Sobol’s sensitivity analysis is one of the most powerful techniques [29]. Using Sobol’s method, we compute the first-order indices to measure the contribution from individual parameters and total-order indices which include all higher-order interactions. Both first-a nd t otal-order i ndices a re p ositive n umbers ( total-order sensitivity indices are greater than the first-order sensitivity indices). Furthermore, in many settings, parameters with sensitivity indices greater than 0.05 are considered significant [29].

The outcome of interest for the GSA is the fraction of individuals in the hospitalized and asymptomatic classes for both strains at the equilibrium, that is, 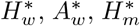 , and 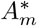 . The ranges used for the parameter values were obtained from past studies on SARS-CoV-2 and are summarized in Table 1. The implementation used the open-source library SALib [30]. The results are presented in Figure 4 and are based on 10^5^ model evaluations. In Figure 4 the left column shows the first- and total-order sensitivity indices whereas the right column shows a histogram of the values of the hospitalized and asymptomatic classes for both strains at the equilibrium. For 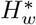 (see the first row in Figure 4), the results indicate that the transmission rates *β*_*w*_ and *β*_*m*_ are the most relevant parameters as their first-order indices above 0.1. Nevertheless, considering all higher-order interactions the mutant recovery rate *γ*_*m*_, the factors *ϕ*_*w*_ and *ϕ*_*m*_, and the wild-type disease-severity ratio *η*_*w*_ are all relevant parameters with total-order indices above 0.1. The histogram for 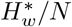 presents a right-skewed distribution which shows that for the majority of the parameter values the number of people in the class *H*_*w*_ will tend to zero and can be as high 5% of the population. It is important to remark that the extinction of the infection in the class *H*_*w*_ might be due to the total eradication of the epidemic but also because the mutant overcomes the wild-type virus. Similar dynamics are observed for 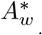 (see the second row in Figure 4) but in addition to *β*_*w*_, and *β*_*m*_, the wild-type disease severity ratio *η*_*w*_ is now also a significant parameter with a first-order index above 0.05 and is, therefore, the third most influential parameter for 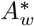

**Table 1:** Parameters for the 2-strain SHAR model (4). WT: wild-type, MT: mutant type, d: days.

| Parameter | Mean | Range | Units | Source |
| --- | --- | --- | --- | --- |
| $\gamma_w$ recovery rate WT | 1/7 | [1/8, 1/3] | d<sup>-1</sup> | [31] |
| $\gamma_m$ recovery rate MT | 1/7 | [1/10, 1/4] | d<sup>-1</sup> | [7, 9] |
| $1/\alpha$ duration of natural immunity | 180 | [0, 365] | d | [32] |
| $\beta_w$ transmission rate WT | 0.2 | [0.1, 1.38] | d<sup>-1</sup> | [7, 9] |
| $\phi_w$ change of infectivity of $A_w$ versus $H_w$ | 1.2 | [0.9, 2.5] | 1 | [21, 22, 25] |
| $\beta_m$ transmission rate MT | 0.4 | [0.1, 1.8] | d<sup>-1</sup> | [7, 9] |
| $\phi_m$ change of infectivity of $A_m$ versus $H_m$ | 1.6 | [1.0, 3] | 1 | assumed |
| $\eta_w$ hospitalized fraction for WT infection | 0.4 | [0.05, 0.60] | 1 | [18] |
| $\eta_m$ hospitalized fraction for MT infection | 0.1 | [0.05, 0.30] | 1 | assumed |
| $\epsilon$ fraction of infections that mutate | $10^{-4}$ | $[10^{-6}, 10^{-3}]$ | 1 | assumed |
| $\rho$ import parameter | $10^{-5}$ | $[10^{-7}, 10^{-3}]$ | 1 | [21, 22, 25] |

**Figure 4.**
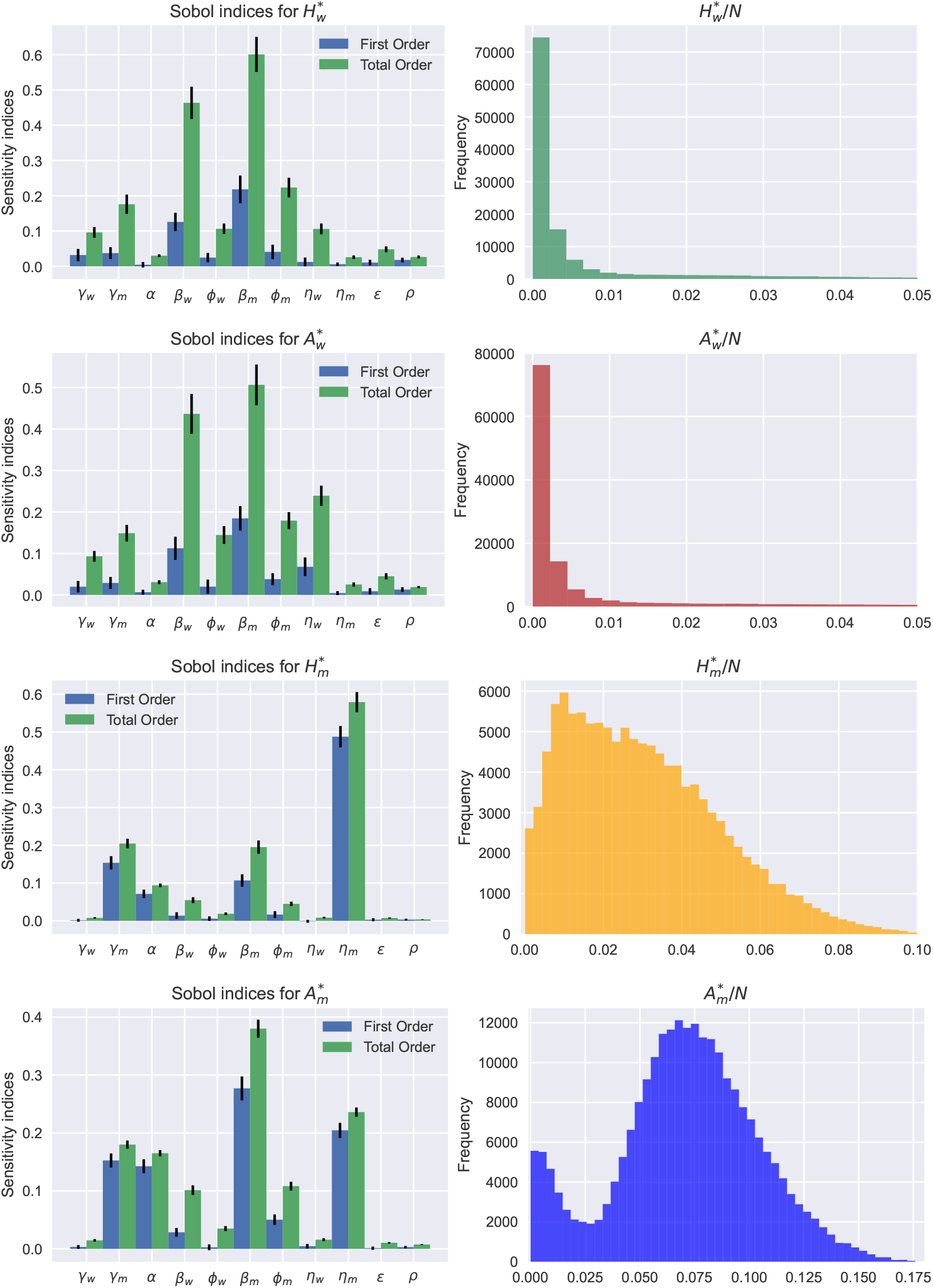
(Left column) First and total order Sobol sensitivity indices. The vertical black lines represent 95% confidence intervals. (Right column) Histograms for the infectious classes at the endemic equilibrium. The total population is fixed as *N* = 10^5^. The ranges explored for the parameters are summarized in Table 1.

For 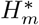 (see the third row in Figure 4), the results indicate that the mutant recovery rate *γ*_*m*_, the loss of immunity *α*, the mutant transmission rate *β*_*m*_, and the mutant hospitalized fraction *η*_*m*_ are the significant parameters, in which *η*_*m*_ is by far the dominant parameter with a first-order sensitivity index being 0.49. The histogram for 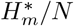 no longer indicates that extinction is the most probable outcome.

Instead, the percentage of people at the equilibrium for the class 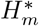 is expected to be between 1 − 8%. For 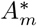 (see the fourth row in Figure 4), the simulations show that *γ*_*m*_, *α, β*_*m*_, *ϕ*_*m*_, and *η*_*m*_ are the significant parameters. The histogram for 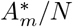 follows a bimodal distribution. The first peak shows a relatively high probability of extinction. However, the second peak is way higher and shows that the most frequent value for 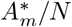 is around 7.5% of the population. Yet the maximum value for 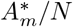 is close to 18% of the population which is almost double the highest value reached by 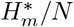 . This larger range for 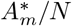 can be explained by the assumption that the mutant generates, on average, more asymptomatic than symptomatic infections. Observe that for the wild-type infectious classes, the total-order indices are much larger than the first-order indices. This suggests that higher-order interactions among several parameter values dominate wild-type dynamics and hence controlling one or two parameters does not guarantee the successful reduction of wild-type prevalence.

### 2.3 Reaction scheme and master equation of the two-strain SHAR model with import

The deterministic model described in the previous section was obtained via the mean-field approximation of the stochastic system described by the following set of reactions. First, we have four reactions related to the infection process via the import *ρ*,

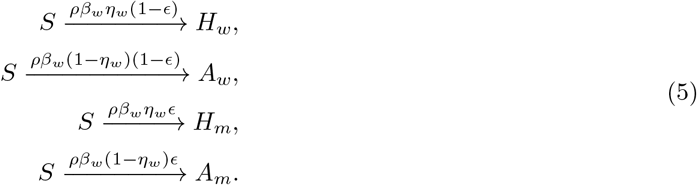

The next reactions correspond to infections after a successful contact of a susceptible with an individual in the *H*_*w*_ class,

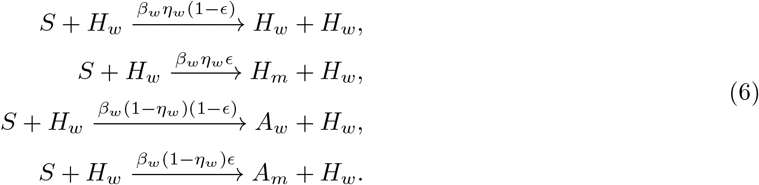

Likewise, we have four reactions corresponding to infections caused by individuals in the *A*_*w*_ class,

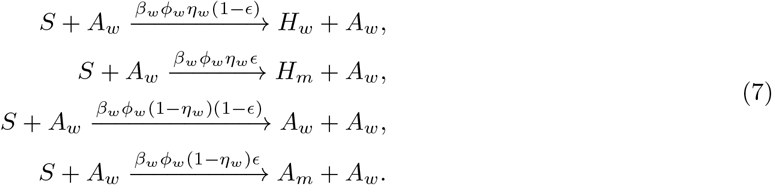

The infections caused by the mutant-type hospitalized infected individuals are,

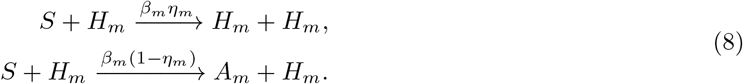

Whereas the infections caused by the mutant-type asymptomatic infected individuals are,

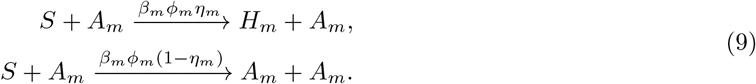

Last, recovery and loss of immunity are described by the next reactions

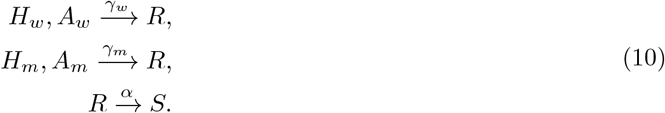

The two-strain stochastic *SHAR* epidemic model with import is modelled as a time-continuous Markov process to capture population noise. Defining the densities of all state variables as *x*_1_ := *S/N* , *x*_2_ := *H*_*w*_*/N* , *x*_3_ := *A*_*w*_*/N* , *x*_4_ := *H*_*m*_*/N* , *x*_5_ := *A*_*m*_*/N* , *x*_6_ := *R/N* , and hence the state vector for the densities *x* := (*x*_1_, *x*_2_, … , *x*_6_)^*tr*^, the master equation can be formulated in a generic form [33, 34]. Within this framework, the evolution of the probability *p*(*x, t*) that the system has a particular composition as a function of time as

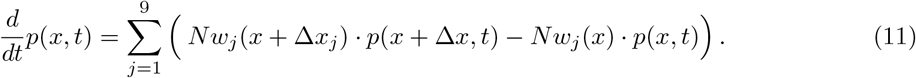

The full expression for the master equation is given in Appendix A. The transitions *w*_*j*_(*x*) (*j* = 1, 2, … , 9) describe the reactions (5)-(10) and 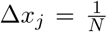 is a small deviation from the densities state vector *x* where *r*_*j*_ (*j* = 1, 2, … , 9) are shifting vectors. The explicit form for the transitions and shifting vectors is as follows:

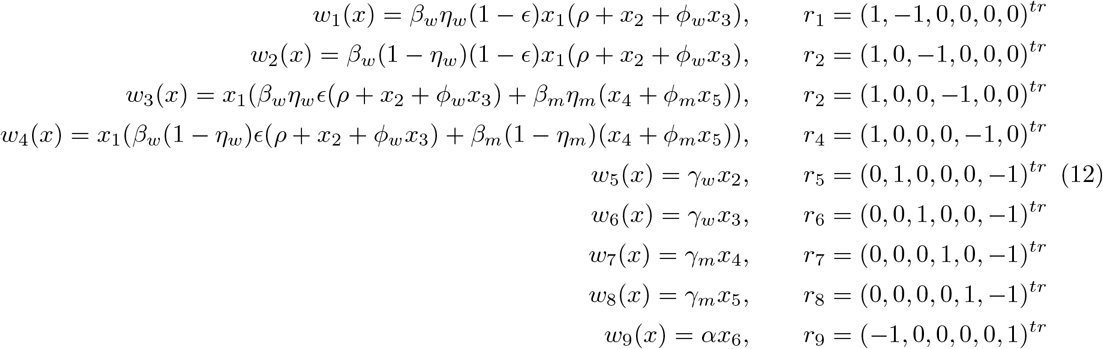

From the two-strain SHAR model given as a master equation, we obtain realizations of the stochastic process via the classical Doob–Gillespie algorithm which provides exact simulations of possible trajectories of the master equation by using standard Monte Carlo techniques [35]. The algorithm is implemented in StochSS [36].

The strain-specific reproduction numbers for system (4) are given by the following expressions:

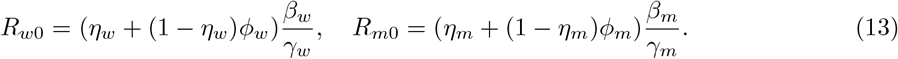

The above reproduction numbers quantify the strain-specific average number of secondary infections that a typical infectious individual generates in a population and are a proxy to approximate variant fitness at the between-host level [12]. All the VOCs that emerged during the COVID-19 pandemic have evolved to maximize their reproduction numbers and hence spread more efficiently than previous variants at the population level [3].

Different pathways can increase the fitness or reproductive success of an invading variant. For instance, increasing transmissibility is a well-known evolutionary process of fitness maximization [4]. The Alpha variant detected at the end of 2020 presented a significant increase in transmissibility over previous SARS-CoV-2 lineages. Later, Beta, Gamma, Delta, and Omicron also presented transmission advantages over preceding variants and were 25-100% more transmissible than the original Wuhan strain [37, 3, 4]. Hence, for the two-strain *SHAR* model is plausible to assume that the mutant has a higher reproduction number. One can get *R*_*w*0_ < *R*_*m*0_ assuming that the mutant has a higher baseline transmission rate, that is, *β*_*w*_ ≤ *β*_*m*_. Since we are considering that asymptomatic and symptomatically infectious individuals have different transmissibility, the reproduction numbers can also be affected by the relative severity of SARS-CoV-2 variants. In particular, assuming that due to higher levels of mobility infectious individuals with mild symptoms might transmit more than people with severe disease then a mutant variant can increase its fitness generating more asymptomatic infections (infections with mild symptoms). For most cases, when a virus causes high fatalities in hosts its transmissibility would be severely limited reducing its probability to survive [38]. Nevertheless, determining the relative severity of SARS-CoV-2 variants is a challenging task, and more work is needed to understand how their virulence evolves [3]. For instance, the variants Alpha and Delta showed greater severity and lethality than their predecessors, whereas Omicron exhibited lower severity and lethality as well as exceptionally high transmissibility [38].

Figure 5 presents an ensemble of stochastic realizations that shows how changes in the mutant variant disease severity ratio impact the population-level prevalence while other parameters are equal for both strains. The disease severity ratio is assumed equal to *η*_*w*_ = 0.4 for the wild-type. Whereas for the mutant it is assumed *η*_*m*_ = 0.2 (first row) and *η*_*w*_ = 0.1 (second row). In Figure 5 (and the subsequent figures) the transparent thin lines correspond to stochastic realizations, whereas the respective bold lines represent the mean-field solutions. The initial condition for the wild-type infectious classes is assumed close to its endemic equilibrium value and for the mutant is assumed close to zero. Since the factors are assumed to satisfy *ϕ*_*w*_ = *ϕ*_*m*_ > 1, the mutant total prevalence overcomes the wild-type due to a relatively small reduction in disease severity. Nevertheless, even if the total prevalence of the infection and the number of people in the asymptomatic class are higher for the mutant, the hospitalizations might still be bigger for the wild-type if the difference in disease severity is not big enough (see the first row in Figure 5). On the other hand, as shown by Omicron, even if a new variant is considerably less virulent, with a significant increase in transmissibility it can lead to more hospitalizations and deaths than its predecessors since it can infect a huge part of the population (see the second row in Figure 5).

**Figure 5.**
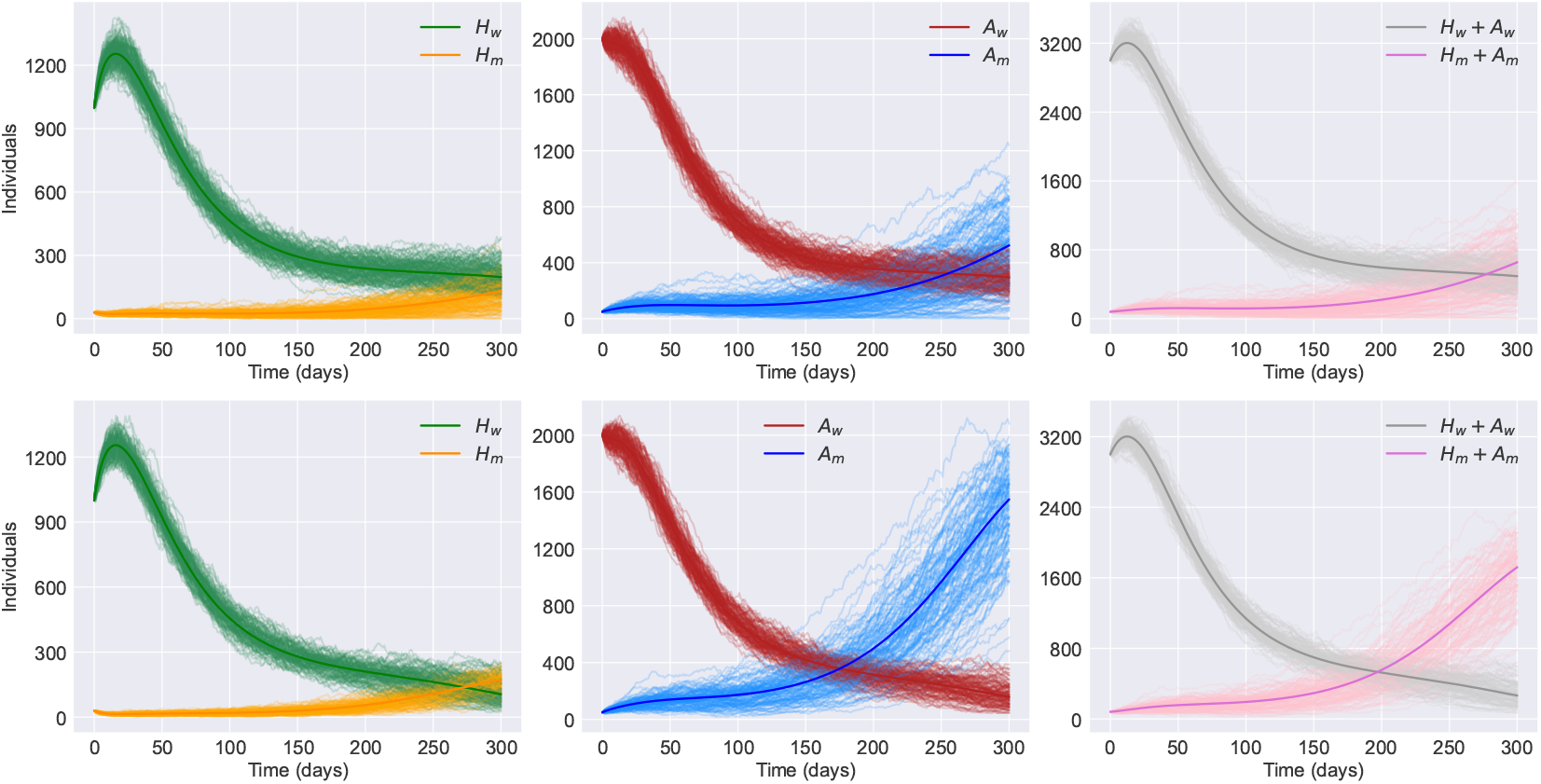
Infected classes for the two-strain stochastic *SHAR* model for *η*_*m*_ = 0.2 (first row) and *η*_*m*_ = 0.1 (second row). The disease-severity ratio for the wild-type is *η*_*w*_ = 0.4 and the rest of the strain-specific parameters are assumed equal for both strains: *γ*_*w*_ = *γ*_*m*_ = 1*/*7, *β*_*w*_ = *β*_*m*_ = 0.8∗ 1*/*7, *ϕ*_*w*_ = *ϕ*_*m*_ = 1.6. The total population is fixed as *N* = 10^5^. Other parameters are fixed with the mean values in Table 1.

The value of the reproduction numbers (13) can also increase if the infectious period is longer so the duration of infectiousness is also an evolvable trait [4]. Figure 6 shows the role of the mean duration of the infectious period on the spread of the infection at the population level. Figure 6 assumes that the wild-type mean infectious period is 1*/γ*_*w*_ = 7 days and for the mutant is 1*/γ*_*m*_ = 8 days while the rest of the parameters are the same for both variants. As a consequence of this one-day increase in the mean duration of the infectious period, the mutant can easily prevail over the wild-type. These results agree with the previous outcomes of the GSA where the recovery rates are highly influential parameters. Hence, a virus with prolonged infectiousness may have a higher likelihood of transmitting to new hosts, as the infected individual remains contagious for an extended period. This can lead to increased opportunities for the virus to spread within a population [4].

**Figure 6.**
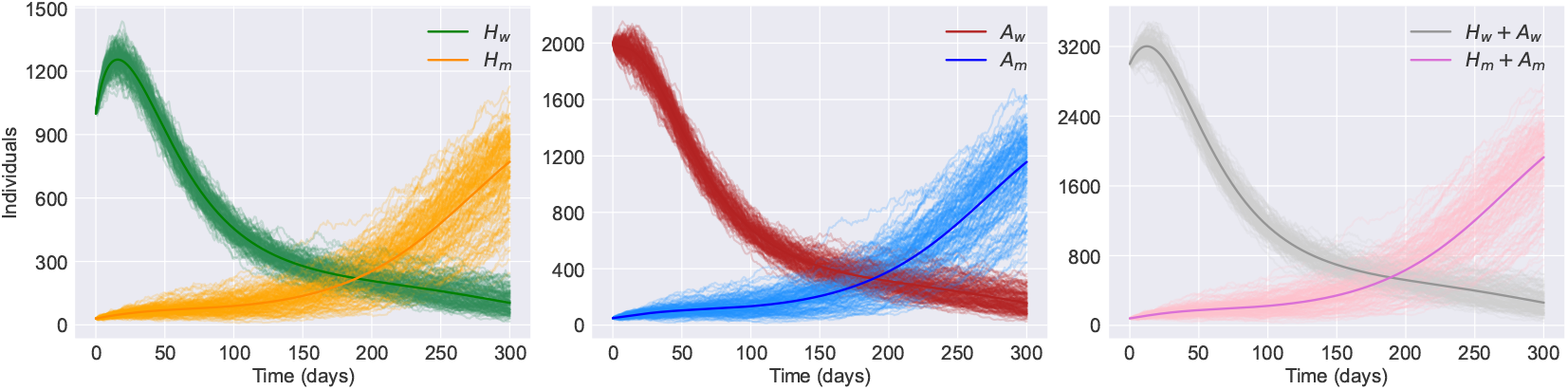
Infected classes for the two-strain stochastic *SHAR* model. The wild-type recovery rate is *γ*_*w*_ = 1*/*7 whereas for the mutant *γ*_*m*_ = 1*/*8. The rest of the strain-specific parameters are assumed equal for both strains: *η*_*w*_ = *η*_*m*_ = 0.4, *β*_*w*_ = *β*_*m*_ = 1*/*7, *ϕ*_*w*_ = *ϕ*_*m*_ = 1.2. The total population is fixed as *N* = 10^5^. Other parameters are fixed with the mean values in Table 1.

## 3 Discussion

Aiming to understand better the emergence, coexistence, and spread of SARS-CoV-2 variants, we developed a two-strain (wild-type and mutant-type) stochastic model that builds upon the classical *SIR* (susceptible-infectious-recovered) model. The model follows a *SHAR* (susceptible-hospitalized-asypmtomatic-recovered) structure and accounts for multiple factors, including asymptomatic transmission, mutations, spillover events, and the potential importation of diseases from external sources into the population of interest. First, we investigated the one-strain deterministic version of the *SHAR* model. We showed that with a positive import *ρ* > 0, the model does not admit a disease-free equilibrium and hence the complete eradication of the disease is not possible. In the absence of disease importation, the one-strain *SHAR* model follows traditional threshold behaviour, showing a stable disease-free equilibrium for *R*_0_ < 1 and a stable endemic equilibrium for *R*_0_ > 1. Furthermore, since the *SHAR* model considers asymptomatic transmission, the basic reproduction number *R*_0_ is the weighted average of the secondary infections caused by the hospitalized and asymptomatic classes. Therefore, even if the *SIR* sub-system is below criticality, considering that the contact rate of the asymptomatic class is significantly higher than the one for the hospitalized class allows the *SHAR* model to reach a supercritical regime (see Figures 1-3). This underscores the role of asymptomatic carriers in COVID-19 spread and emphasizes the need for extensive testing and contact tracing efforts.

Due to the system’s high nonlinearity, it was not possible to perform an analytical analysis for the equilibria of the two-strain deterministic *SHAR* model. Hence, we performed a global sensitivity analysis (GSA) to understand the long-term dynamics of the two-strain model and to measure the individual importance of each parameter as well as their joint effect on model outcomes (see Figure 4). The outcome of interest for the GSA is the fraction of individuals in the hospitalized and asymptomatic classes, 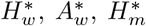 , and 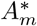 , for both strains at the equilibrium. The histograms for 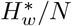 and 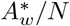 exhibit a right-skewed distribution, indicating that a significant portion of individuals in these classes tends towards zero, with a potential peak at around 5% of the population. The decline in the wild-type infectious classes can be attributed to the complete eradication of the epidemic, although a more probable explanation is the increased transmissibility of the mutant variant over the wild-type virus. Analysis of the Sobol sensitivity indices reveals that the transmission rates *β*_*w*_ and *β*_*m*_ play pivotal roles in determining the dynamics of 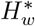 and 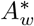 . Notably, the total-order sensitivity indices outweigh the first-order indices for the wild-type infectious classes, suggesting that complex interactions between various parameters significantly influence wild-type dynamics. Hence controlling one or two parameters does not guarantee the successful reduction of wild-type prevalence. The histogram for 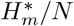 suggests that the equilibrium percentage for this class typically falls within the range of 1-8%. Additionally, the mutant-type disease severity ratio *η*_*m*_ emerges as the most influential parameter, with a first-order sensitivity index approaching 0.5. In contrast, the histogram for 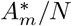 displays a bimodal distribution, with one peak indicating a considerable probability of extinction and another peak showing that the prevalent value for 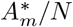 can be in the order of magnitude of 10% of the population. However, the upper limit for 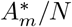 is almost twice the peak value observed for 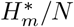 . This broader range for 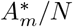 may be attributed to the hypothesis that the mutant variant generates a higher proportion of asymptomatic infections compared to symptomatic cases.

Using the master equation formalism and stochastic realizations via the Doob-Gillespie algorithm, we explored different pathways that may affect the reproductive success of an invading variant. The simulations suggest that contrary to popular belief variants with lower severity are likely to spread more rapidly compared to more severe variants (see Figure 5). However, in agreement with [39], the explanation behind this phenomenon is that variants that cause more severe symptoms might prompt affected individuals to seek medical care sooner, potentially leading to earlier identification and isolation of cases. On the other hand, if a variant causes milder symptoms or is predominantly asymptomatic, individuals may unknowingly transmit the virus to a larger number of people, making it challenging to contain the infection [26, 10]. Furthermore, even if emerging variants are less pathogenic than the wild type, once they become dominant in a population, they might lead to an increase in hospitalizations and potential deaths since the mutant carriers can infect a significantly larger segment of the population [38]. The simulations also show that extended infectiousness of SARS-CoV-2 variants increases considerably the value of the reproduction number and this increase can result in rapid and sustained community spread (see Figure 6) [4]. In other words, a longer period of infectiousness means that individuals infected with these variants can shed the virus and transmit it to others for an extended duration compared to variants with shorter infectious periods.

The primary aim of this study was to explore various scenarios of disease spread to better understand the factors influencing variants’ fitness and evolution. The proposed model describes well the interplay between viral characteristics and transmission dynamics, providing a foundation for further investigation into mitigating strategies and public health interventions. Nevertheless, for future work, accounting for mass vaccination campaigns, particularly concerning vaccine escape [40, 41], and considering the possibility of multiple epidemic waves or seasonal infection patterns could be considered to improve modelling accuracy.

## Data Availability

We do not analyse or generate any datasets, because our work proceeds within a theoretical and mathematical approach.

## Ethics

This work did not require ethical approval from a human subject or animal welfare committee.

## Declaration of AI use

We have not used AI-assisted technologies in creating this article.

## Authors contributions

**Fernando Saldaña**: Conceptualization, Methodology, Software, Formal analysis, Writing - Original draft, Writing - Review & Editing. **Nico Stollenwerk**: Conceptualization, Methodology, Formal analysis, Supervision. **Maíra Aguiar**: Conceptualization, Methodology, Formal analysis, Writing - Review & Editing, Supervision, Funding acquisition.

## Appendix A Master equation for the two-strain SHAR model

Let *X* = (*S, H*_*w*_, *A*_*w*_, *H*_*m*_, *A*_*m*_, *R*)^*tr*^ be the vector of states, then the temporal dynamics for the probability *p*(*X, t*) of having at time *t* an integer number of *S* susceptible, *H*_*w*_ hospitalized and *A*_*w*_ asymptomatically infected with the wild-type, *H*_*m*_ hospitalized and *A*_*m*_ asymptomatically infected with the mutant, and *R* recovered can be described by the following master equation

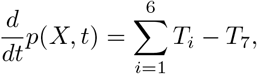

where the terms *T*_1_, *T*_2_, … , *T*_6_ are functions describing the reactions (5)-(10), respectively, and are defined next: The term *T*_7_ in the master equation describes the outflow of the system

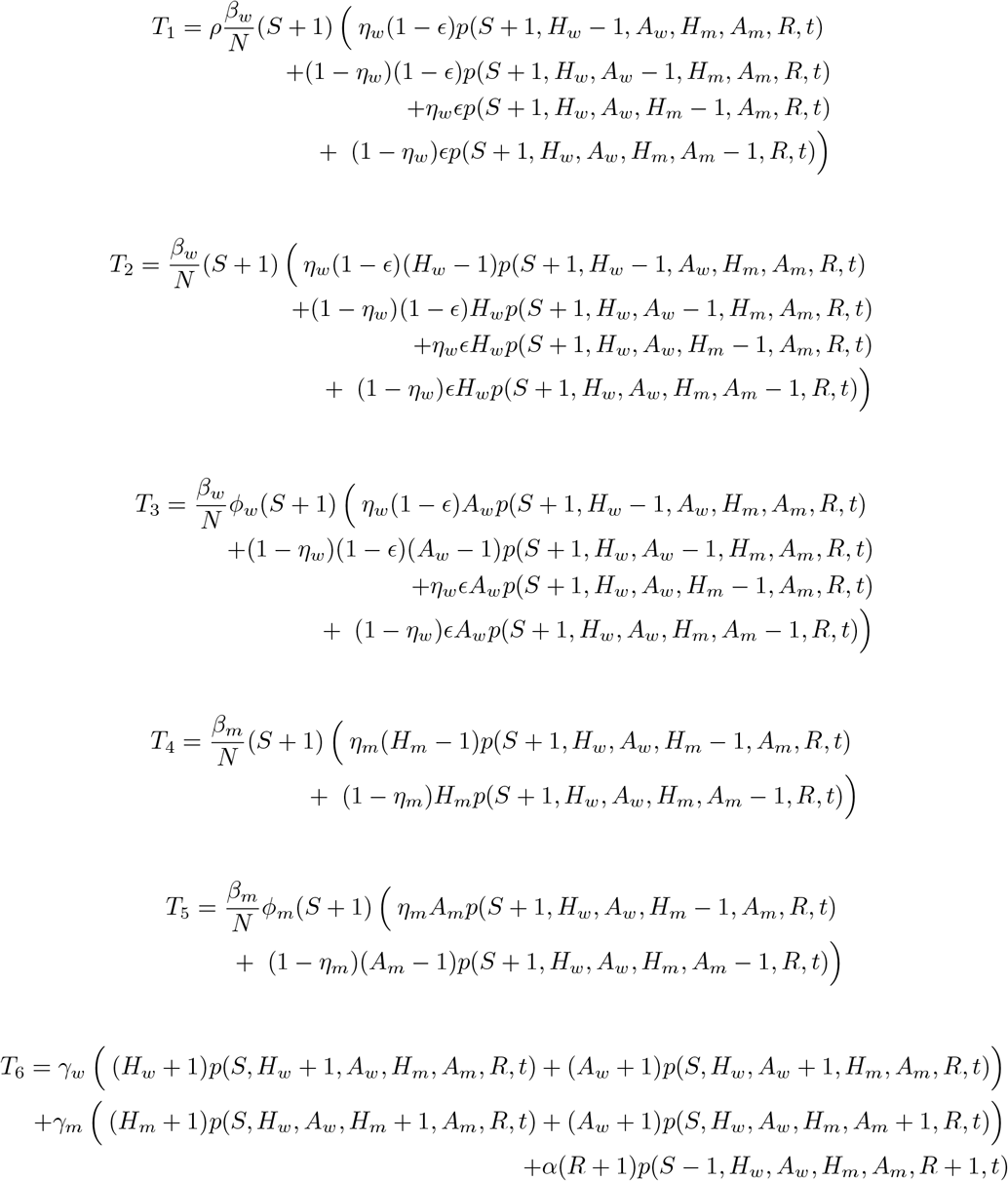

The term T_7_ in the master equation describes the outflow of the system

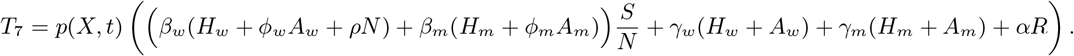

